# Adequate antenatal care visits are associated with a reduction in the odds of low birth weight in Zambia: evidence from the Zambia demographic and health survey

**DOI:** 10.64898/2026.01.10.26343851

**Authors:** Teebeny Zulu, Gracious Ngosa, George Zulu, Herbert Luhanga, Choolwe Jacobs

## Abstract

**Background:** Low birth weight (LBW) remains a major public health challenge in low and middle-income countries and is associated with increased risks of neonatal mortality, impaired growth, and adverse developmental outcomes. Antenatal care (ANC) provides essential interventions that support maternal health and fetal growth; however, many women do not achieve the recommended number of ANC visits. This study is therefore aimed at providing evidence of the protective effect of adequate ANC against LBW in the Zambian context.

**Methods:** We conducted a cross-sectional analysis of data from the 2001-2024 Zambia Demographic and Health Survey (ZDHS). The study included non-anemic women aged 15–49 years with a recent singleton live birth and available birth weight information. Low birth weight was defined as birth weight <2500 g. Antenatal care attendance was categorized as 0–3, 4–7, and ≥8 visits. Choropleth maps and trendlines were used to show the spatial and temporal variation of LBW. Survey-weighted descriptive statistics, logistic regression models and predicted probability were used to estimate crude and adjusted associations between ANC visits and LBW while accounting for the complex survey design. All analysis were performed in STATA version 17.

**Results:** A total of 3644 non-anemic women with recent singleton live births were included in the analysis. The overall prevalence of LBW was 8.7%. The prevalence of LBW was higher among women who attended 0–3 ANC visits compared with those who attended 4–7 visits or ≥8 visits. In adjusted analyses, attending 4–7 visits (AOR; 0.57; 95% CI: 0.42–0.78) and ≥8 visits (AOR 0.51; 95% CI: 0.27–0.99) was associated with lower odds of LBW compared with 0–3 visits. The probability of LBW decreased with an increase in the numbers of ANC visits.

**Conclusion:** Low birth weight remains a significant public health concern in Zambia. Higher ANC attendance was associated with lower odds of LBW, supporting the importance of achieving adequate and sustained ANC during pregnancy. Strengthening strategies to promote early initiation and continuity of ANC, alongside addressing broader socioeconomic and nutritional determinants, may contribute to improved birth outcomes in Zambia.

## Introduction

Low birth weight is defined by the World Health Organization (WHO) as weight at birth less than 2500g [1]. Low birth weight continues to be a significant public health problem globally and is associated with a range of both short- and long-term consequences. Overall, it is estimated that 15% to 20% of all births worldwide are LBW, representing more than 20 million births a year [1]. Reducing LBW is a key global public health priority and forms part of the Global Nutrition Targets for 2025, which aim to achieve a 30% reduction in LBW worldwide [1]. Infants born with LBW are at substantially higher risk of neonatal mortality, childhood morbidity, impaired growth, and adverse developmental outcomes compared with infants born at normal birth weight [2–4].

Low birth weight is a multifactorial phenomenon shaped by interrelated maternal, child, household, and health system factors. Previous studies have found low education, unemployment, poverty, and rural residence to be associated with an increased risk of LBW [5,6]. Maternal undernutrition, anemia, micronutrient, and inadequate ANC visits (< 4 ANC visits) have also been linked to increased risk of LBW [7–10]. Pregnancy and obstetric related factors such as very low or high parity, short birth intervals, and multiple gestations are also associated with increased odds of LBW [6,8]. Maternal infections, unhealthy maternal behaviors, and environmental exposures, including smoking, alcohol use, indoor air pollution, and poor sanitation, further exacerbate the risk of LBW [7]. Additionally, child-related factors such as female sex and broader contextual factors, including limited access to health facilities and regional disparities in service availability, have been shown to influence the occurrence of low birth weight [11,12].

Antenatal care is a core component of maternal health services and plays a critical role in improving pregnancy outcomes. Through ANC, pregnant women receive essential interventions, including nutritional counselling and supplementation, screening and management of infections and pregnancy-related complications, and preventive services aimed at improving maternal and fetal health. Until 2016, the World Health Organization recommended a minimum of four ANC visits during pregnancy under the focused antenatal care model; this recommendation was subsequently revised to eight or more ANC contacts to improve maternal and perinatal outcomes and women’s experience of care [9,13]. Adequate ANC attendance (≥ 4 ANC visits) may reduce the risk of LBW by improving maternal nutritional status and facilitating early detection and management of pregnancy-related conditions. In malaria-endemic settings, adequate ANC provides a platform for the delivery of intermittent preventive treatment for malaria during pregnancy, which is administered during the second and third trimesters and has been shown to reduce the risk of LBW [14,15].

In Zambia, LBW remains a persistent public health concern despite improvements in maternal and child health indicators. The country continues to experience a substantial burden of malaria and maternal undernutrition, particularly in rural and socioeconomically disadvantaged populations, both of which are established risk factors for LBW [6,16]. Although overall adequate ANC coverage in Zambia is relatively high, disparities in ANC utilization persist across geographic and socioeconomic groups, potentially limiting the protective benefits of ANC for some women. Evidence on the relationship between ANC attendance and LBW in Zambia remains limited and is largely based on older survey rounds or simplified dichotomizations of ANC utilization. Moreover, few studies have examined LBW prevalence and adjusted associations across ANC visit categories that reflect both the former four-visit recommendation and the current eight-contact guideline using recent nationally representative data. This study is therefore aimed at establishing the effect of adequate ANC on LBW in Zambia.

## 2. Methods

### 2.1 Study area and setting

This study was conducted using nationally representative sample from Zambia, a landlocked country in southern Africa and part of sub-Saharan Africa (SSA). The analysis covered all ten provinces of the country. Zambia’s total population was estimated at approximately 21.9 million in 2025, with an annual population growth rate of about 2.8% [17,18]. The population is predominantly rural, with approximately 53% of Zambians residing in rural areas, while the remainder live in urban settings [18]. Of these about 30% are women of reproductive age (15-49 years), reflecting persistently high fertility and demographic patterns typical of many countries in SSA [18].

### 2.2 Study design, data source and study population

This study employed a cross-sectional design using data from the 2001-2024 ZDHS. Datasets for each survey year (2001-2024) were used to generate trendlines and choropleth maps while the 2024 dataset was used for inferential statistics. The ZDHS is a nationally representative household survey implemented by the Zambia Statistics Agency in collaboration with international partners, including ICF International and the United States Agency for International Development (USAID). The survey uses a two-stage stratified sampling design, in which enumeration areas (EAs) are selected in the first stage, followed by the systematic selection of households in the second stage. In selected households, all eligible women aged 15–49 years who were usual residents or who slept in the household the night before the survey were eligible for interview. Information was collected on demographic characteristics, reproductive health, antenatal care utilisation, and birth outcomes. For the present analysis, the study population was restricted to non-anaemic singleton live births with recorded birth weight in the reference period preceding the survey. We only included last born and singleton live births from mothers who were non-anaemic during pregnancy because these are considered high risky pregnancies. Multiple pregnancy is a known risk factor of LBW and is likely to mask the effect of other variables [19,20]. We chose to include only last-born children for several reasons: first, information about the most recent birth is likely to be more accurate and less subject to recall bias compared to earlier births; second, including more than one birth from one mother would have included maternal and household variables more than once, potentially biasing the results towards families with multiple children, and third, the circumstances surrounding the last birth are more likely to reflect the current socioeconomic status of the household [21]. We further chose to exclude anaemic mothers because they have more prescribed ANC visits than non-anaemic mothers hence this could also mask the effect of having adequate ANC visits on LBW. We further excluded missing observations in low birthweight and more than 50% missing in key covariates, thus a total of 3690 live births were included in the analysis, representing a weighted population of 3644 births.

### 2.3 Study variables

#### 2.3.1 The outcome variable

The outcome in this study was LBW. Birth weights were retrieved based on the mother’s recall and/or from a birth card. The continuous birth weight variable was then dichotomized as LBW if < 2500 g and ‘non-LBW’ if otherwise, in accordance with the global standard [22,23].

#### 2.3.2 The primary exposure variable

The main exposure variable was number of antenatal care visits. This variable was collected as a count in 2024 ZDHS. This variable was categorized into 0-3 visits, 4-7 visits and 8 or more visits. This categorization was chosen to reflect the old WHO minimum recommendation of 4 or more (adequate) ANC visits and the current 8 or more ANC visits [9,24]. The reference category was merged into 0-3 because there were very few mothers with 0 ANC visits. Since categorizing continuous variables leads to loss of information, we also considered ANC visits as counts to show the dose response of increasing the number of ANC on the predicted probability of LBW.

#### 2.3.3 Secondary exposure variables

The effect of ANC visits on low birthweight was adjusted for the following variables; number of children ever born (*count*), maternal employment status (*not employed/employed*), sex of a child (*male/female*), type of toilet facility (*improved/unimproved*), maternal stunting (*stunted/not stunted*), place of residence (*rural/urban*) and province (*Central, Copperbelt, Eastern, Luapula, Lusaka, Muchinga, Northern, North-Western, Southern, Western*)

#### 2.3.4 Statistical analysis

Data analysis was performed in STATA version 17. We reported survey weighted frequencies and percentages for categorical variables. The median and interquartile range was reported for skewed continuous variables. We reported the prevalence of LBW by categories of adequate ANC and other socio-demographic characteristics. Choropleth maps and trendlines generated in R statistical software, were used to show the spatial and temporal trends of the survey weighted prevalence of LBW. Unadjusted and adjusted logistic regression was performed to determine the effect of ordinal ANC visits on LBW using the 2024 ZDHS. Adjusted logistic regression was fitted with continuous number of ANC visits to obtain marginal predicted probabilities of the effect of ANC on LBW. These probabilities were plotted with confidence intervals to visually show the dose response of ANC on LBW. The analysis accounted for the complex survey design of the 2024 ZDHS by applying sampling weights, clustering, and stratification. Sampling weights were derived from the women’s individual sample weight (v005), which was rescaled by dividing by 1000000, while primary sampling units and sampling strata were specified using the cluster variable (v021) and the stratification variable (v022), respectively.

### 2.7 Ethical considerations

The ICF Institutional Review Board approved the ZDHS data survey protocols with the ICF Project Number: 132989.0. 000.ZM. DHS.02. Permission to use the dataset was acquired from ICF Macro, and the dataset named ZMBR81DTA is available for download at https://www.dhsprogram.com/data. The user carefully adhered to the given guidelines, highlighting the sensitive nature of the information and the significance of not trying to uncover the identity of any household or individual surveyed for the study (maintaining anonymity).

## 3. Results

### 3.1 Overall characteristics of the study population

The analysis included 3644 live births, of which 8.7% were classified as LBW, while 91.3% had normal birth weight (Table 1). In the overall sample, most women (78.5%) attended 4–7 antenatal care (ANC) visits, 16.1% attended 0–3 visits, and 5.4% attended eight or more visits. Slightly more than half of mothers were not employed (52.8%), and infants were almost evenly distributed by sex (50.3% male and 49.7% female). Over half of households had improved toilet facilities (54.8%), while 45.2% used unimproved facilities. The majority of mothers were not stunted (88.5%). The median number of children ever born was 2 (interquartile range [IQR]: 1–4). Geographically, births were most common in Lusaka Province (15.9%), followed by Eastern (13.0%), Copperbelt (12.9%), Southern (12.5%), and Central (12.2%) provinces.

**Table 1:** Distribution of LBW across categories of ANC visits and the sociodemographic characteristics of the respondents.

| Variable | Overall<br>n = 3644 (100%) | Low birth weight |  | p-value |
| --- | --- | --- | --- | --- |
|  |  | No<br>n = 3325 (91.3%) | Yes<br>n = 318 (8.7%) |  |
| **Number of antenatal care visits** |  |  |  |  |
| 0–3 visits | 586 (16.1) | 510 (87.0) | 76 (13.0) |  |
| 4–7 visits | 2860 (78.5) | 2632 (92.0) | 228 (8.0) |  |
| ≥8 visits | 199 (5.4) | 184 (92.6) | 15 (7.4) | 0.0021 |
| **Maternal employment status** |  |  |  |  |
| Not employed | 1924 (52.8) | 1731 (90.0) | 193 (10.0) |  |
| Employed | 1720 (47.2) | 1594 (92.7) | 126 (7.3) | 0.0067 |
| **Sex of the child** |  |  |  |  |
| Male | 1832 (50.3) | 1692 (92.3) | 140 (7.7) |  |
| Female | 1812 (49.7) | 1633 (90.2) | 178 (9.8) | 0.0274 |
| **Type of toilet facility** |  |  |  |  |
| Improved | 1991 (54.8) | 1830 (91.9) | 161 (8.1) |  |
| Unimproved | 1643 (45.2) | 1485 (90.4) | 157 (9.6) | 0.1456 |
| **Maternal stunting status** |  |  |  |  |
| Not stunted | 3226 (88.5) | 2956 (91.6) | 270 (8.4) |  |
| Stunted | 418 (11.5) | 370 (88.4) | 48 (11.6) | 0.0281 |
| **Number of children ever born** | 2 (1–4) | 3 (1–4) | 2 (1–4) | 0.0001 |
| **Province** |  |  |  |  |
| Central | 443 (12.2) | 387 (87.4) | 56 (12.6) |  |
| Copperbelt | 470 (12.9) | 435 (92.6) | 35 (7.4) |  |
| Eastern | 475 (13.0) | 435 (91.6) | 40 (8.4) |  |
| Luapula | 313 (8.6) | 290 (92.8) | 23 (7.2) |  |
| Lusaka | 580 (15.9) | 527 (90.9) | 53 (9.1) |  |
| Muchinga | 183 (5.0) | 167 (91.6) | 15 (8.4) |  |
| Northern | 305 (8.4) | 281 (92.1) | 24 (7.9) |  |
| North Western | 235 (6.5) | 222 (94.4) | 13 (5.6) |  |
| Southern | 457 (12.5) | 413 (90.3) | 44 (9.7) |  |
| Western | 184 (5.1) | 168 (91.2) | 16 (8.8) | 0.1613 |

Survey-weighted chi-square (Rao-Scott) test was used to check for associations between categorical predictors and LBW. Row counts and percentages are presented to show the dose-response relationship. Man-Whitney test was used for a continuous predictor.

### 3.2 National trends of adequate ANC utilization and LBW

At the national level, coverage of at least four ANC visits declined between 2001 and 2013, reaching its lowest level during this period, before increasing substantially in subsequent survey years (Fig 1). In contrast, the prevalence of LBW showed relatively modest variation over time, with a slight decline up to 2018 followed by a small increase in 2024. The recovery in ANC coverage after 2013 coincided with a stabilization of LBW prevalence at comparatively low levels. The observed trends suggest an inverse temporal relationship between ANC utilization and LBW prevalence which could indicate that changes in LBW are likely influenced by multiple factors beyond ANC attendance in isolation.

**Figure 1:**
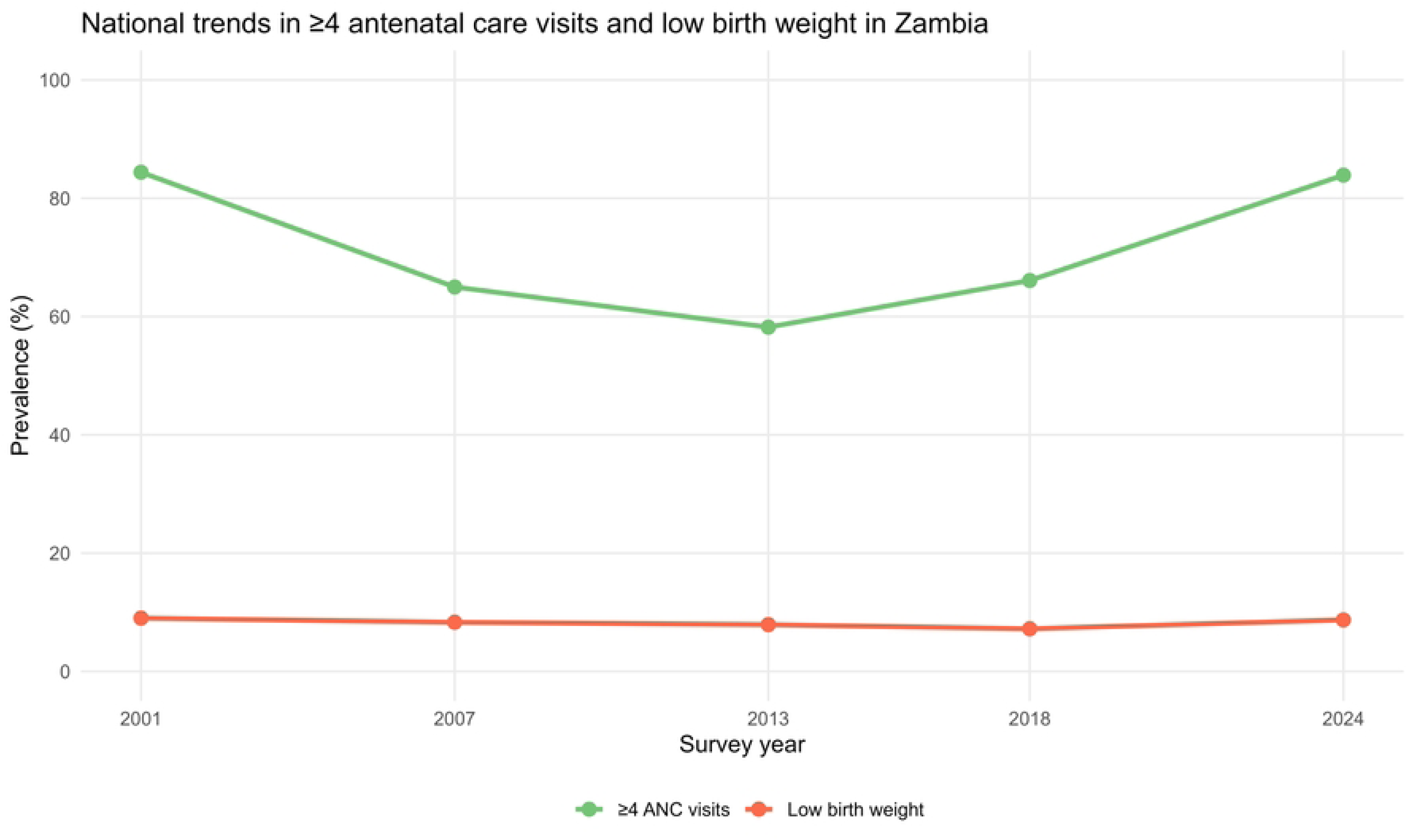
National trends of ≥4 ANC visits and LBW in Zambia from 2001-2024 survey years. The green and red trendlines represents the survey-weighted prevalence of ≥4 ANC visits and LBW respectively.

### 3.2 Spatial distribution of LBW and ANC utilization by province over survey years

Spatial and temporal heterogeneity in the prevalence of LBW across Zambian provinces between 2001 and 2024 exists (Fig 2). In the early survey years (2001 and 2007), higher LBW prevalence was concentrated in provinces such as Luapula, Western, Northern, and parts of Southern Province, indicating persistent geographic inequalities. Although some provinces exhibited reductions in LBW prevalence by 2013, progress was uneven, and by 2018 and 2024 several provinces including Central, Southern, and Lusaka showed stabilisation or renewed increases in LBW prevalence.

**Figure 2:**
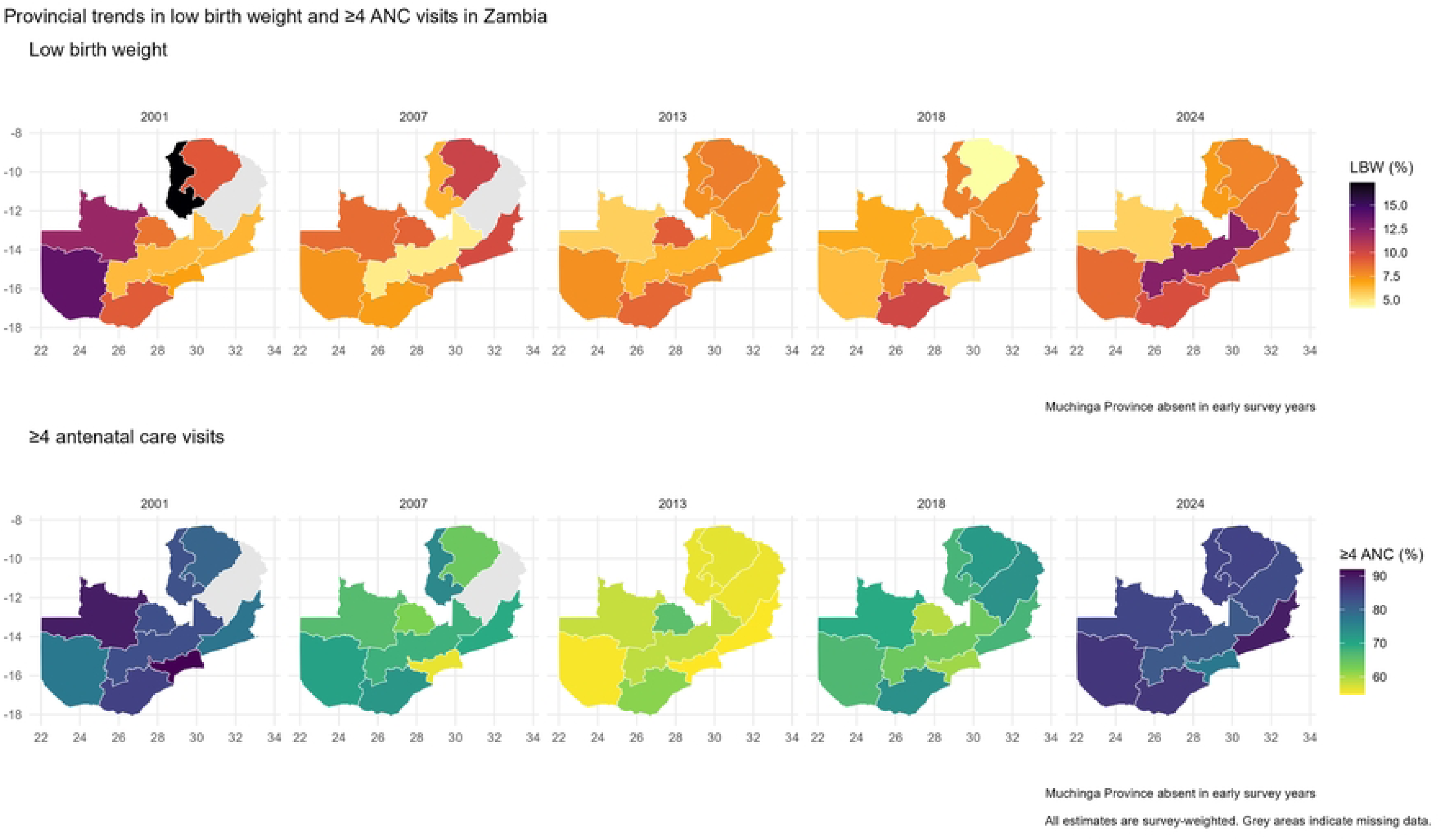
Spatial trends of LBW and ≥4 ANC visits by province over 2001-2024 survey years. Maps showing provincial prevalence of LBW (top row) and coverage of ≥4 ANC visits (bottom row) in Zambia across survey years. Darker shading indicates higher values. All estimates are survey-weighted. Grey areas indicate provinces not observed in a given survey year (Muchinga Province absent in early surveys).

In contrast, coverage of at least four ANC visits increased substantially and consistently over time, with marked convergence across provinces by 2024. While early surveys revealed considerable regional disparities in ANC utilisation, later survey years show high coverage in nearly all provinces, reflecting broad improvements in access to maternal health services. Notably, however, provinces with high ANC coverage in 2024 did not uniformly exhibit low LBW prevalence, indicating a disconnect between service contact and birth outcomes. This divergence suggests that increases in ANC coverage alone may be insufficient to reduce LBW without concurrent improvements in the timing, quality, and content of care, as well as in broader maternal health, nutritional, and socioeconomic conditions

### 3.3 Trends of LBW by province from 2001-2024 ZDHS survey years

Province-specific trends in 4 or more ANC and LBW prevalence varied considerably over time in Zambia from 2001 to 2024 (Fig 3). Across all provinces, coverage of at least 4 ANC visits is consistently high and shows an overall upward trajectory over time, with a noticeable dip around 2007–2013 followed by substantial recovery by 2024. By the most recent survey, nearly all provinces reach very high ANC coverage, indicating widespread access to antenatal services.

**Figure 3:**
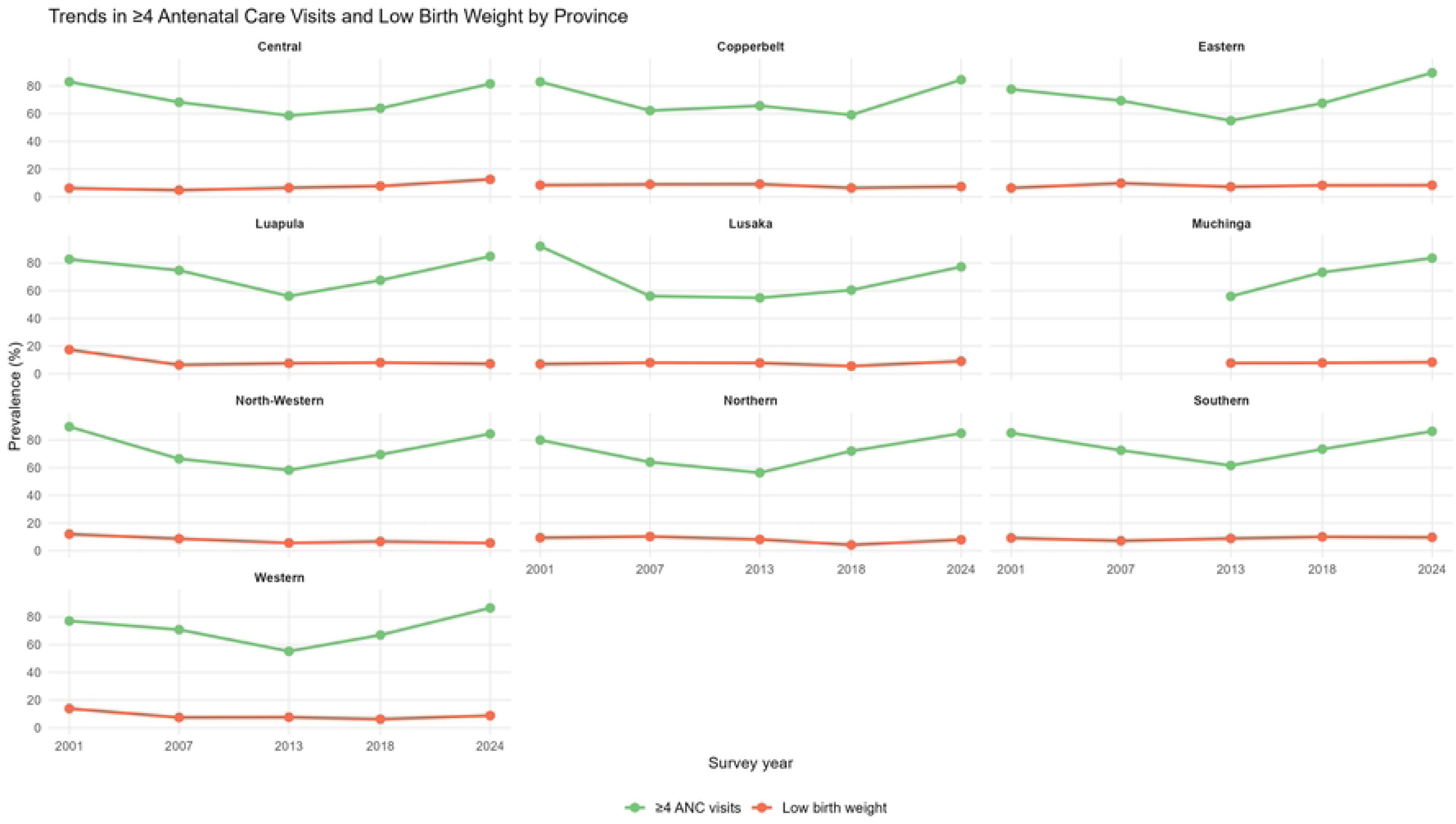
Temporal trends of LBW and ≥4 ANC visits by province over 2001-2024 survey years. Trendlines showing provincial prevalence of LBW (red) and coverage of ≥4 ANC visits (green) in Zambia across 2001-2024 survey years. All estimates are survey-weighted. The trendlines for Muchinga province are truncated since it was created after the 2007 survey.

In contrast, trends in LBW remain persistently low in magnitude but heterogeneous in direction, with most provinces fluctuating within a narrow range below 12%. While several provinces demonstrated declining or stable prevalence between 2001 and 2018, recent increases were observed in Central, Lusaka, Southern, Northern, and Western provinces in 2024. Luapula and North-Western provinces showed sustained reductions following high prevalence in the early 2000s, whereas Muchinga exhibited relatively stable prevalence since its inclusion in the surveys.

### 3.3 Distribution of LBW across ANC categories and sociodemographic characteristics of the respondents

The prevalence of LBW varied significantly by ANC attendance (p = 0.0021). LBW prevalence was highest among mothers who attended 0–3 ANC visits (13.0%), compared with 8.0% among those with 4–7 visits and 7.4% among those with eight or more visits. Furthermore, LBW prevalence also differed by maternal employment status (p = 0.0067), with a higher prevalence among unemployed mothers (10.0%) compared with employed mothers (7.3%). Female infants had a significantly higher prevalence of LBW (9.8%) than male infants (7.7%; p = 0.0274). in addition, LBW prevalence was significantly higher among children born from stunted mothers (11.6%) than among non-stunted children (8.4%) (p = 0.0281). The median number of children ever born differed significantly by LBW status (p = 0.0001), with mothers of LBW infants having a lower median parity (2 children; IQR: 1–4) compared with mothers of normal-birth-weight infants (3 children; IQR: 1–4). Across provinces, LBW prevalence ranged from 5.6% in North Western Province to 12.6% in Central Province; however, these differences were not statistically significant (p = 0.113).

### 3.4 Univariable logistic regression of adequate ANC on LBW

Before adjusting for any confounders, attending 4-7 ANC visits as compared to attending 0-3, was associated with a reduction in the odds of low birthweight by 42% (Table 2). This reduction could be as low as 21% to as high as 57% in Zambia, p-value <0.001. Furthermore, as compared to attending 0-3 ANC visits, attending eight and more ANC visits reduced the odds of low birthweight by 46% (COR = 0.54; 95% CI: 0.28– 1.03; p < 0.062. The number of children ever born was inversely associated with LBW. Each additional birth was associated with a 9% reduction in the odds of LBW (COR = 0.91; 95% CI: 0.84–0.97; p = 0.006). furthermore, infants born to employed mothers had 29% lower odds of LBW compared with those born to unemployed mothers (COR = 0.71; 95% CI: 0.55–0.91; p = 0.007). Female infants had 31% higher odds of being born with LBW compared with male infants (COR = 1.31; 95% CI: 1.03–1.68; p = 0.028). Type of toilet facility was not significantly associated with LBW in crude analysis. Infants from households with unimproved toilet facilities had higher odds of LBW compared with those from households with improved facilities, but this association was not statistically significant (COR = 1.20; 95% CI: 0.94–1.54; p = 0.146). Maternal stunting was significantly associated with LBW. Children from stunted mothers had 43% higher odds of LBW compared with non-stunted children (COR = 1.43; 95% CI: 1.04–1.98; p = 0.029). Infants born in rural areas had higher odds of LBW compared with those born in urban areas (COR = 1.29; 95% CI: 1.00–1.67; p = 0.049). Several provinces showed significantly lower odds of LBW compared with Central Province. Lower odds were observed in Copperbelt (COR = 0.55; p = 0.008), Luapula (COR = 0.54; p = 0.018), Northern (COR = 0.60; p = 0.029), and North Western provinces (COR = 0.41; p = 0.004). Associations for Eastern, Lusaka, Muchinga, Southern, and Western provinces were not statistically significant.

**Table 2.** Crude and adjusted effect of antenatal care visits on LBW in Zambia, ZDHS 2024.

| Variable | COR (95% CI) | p-value | AOR (95% CI) | p-value |
| --- | --- | --- | --- | --- |
| **Number of antenatal care visits** |  |  |  |  |
| 0–3 visits (Inadequate) | Ref |  | Ref |  |
| 4–7 visits | 0.58 (0.43–0.79) | <0.001 | 0.57 (0.42–0.78) | <0.001 |
| ≥8 visits | 0.54 (0.28–1.03) | 0.062 | 0.51 (0.27–0.99) | 0.047 |
| **Number of children ever born** | 0.91 (0.84–0.97) | 0.006 | 0.90 (0.84–0.97) | 0.005 |
| **Employment status** |  |  |  |  |
| Not employed | Ref |  | Ref |  |
| Employed | 0.71 (0.55–0.91) | 0.007 | 0.75 (0.57–0.97) | 0.031 |
| **Sex of the child** |  |  |  |  |
| Male | Ref |  | Ref |  |
| Female | 1.31 (1.03–1.68) | 0.028 | 1.32 (1.03–1.69) | 0.028 |
| **Type of toilet facility** |  |  |  |  |
| Improved | Ref |  | Ref |  |
| Unimproved | 1.20 (0.94–1.54) | 0.146 | 1.15 (0.86–1.53) | 0.349 |
| **Maternal stunting status** |  |  |  |  |
| Not stunted | Ref |  | Ref |  |
| Stunted | 1.43 (1.04–1.98) | 0.029 | 1.49 (1.06–2.10) | 0.021 |
| **Place of residence** |  |  |  |  |
| Urban | Ref |  | Ref |  |
| Rural | 1.29 (1.00–1.67) | 0.049 | 1.33 (0.98–1.81) | 0.071 |
| **Province** |  |  |  |  |
| Central | Ref |  | Ref |  |
| Copperbelt | 0.55 (0.35–0.86) | 0.008 | 0.68 (0.43–1.08) | 0.101 |
| Eastern | 0.64 (0.39–1.03) | 0.065 | 0.59 (0.36–0.96) | 0.036 |
| Luapula | 0.54 (0.32–0.90) | 0.018 | 0.56 (0.34–0.94) | 0.027 |
| Lusaka | 0.69 (0.45–1.06) | 0.094 | 0.84 (0.54–1.30) | 0.430 |
| Muchinga | 0.64 (0.38–1.08) | 0.094 | 0.61 (0.35–1.06) | 0.078 |
| Northern | 0.60 (0.38–0.95) | 0.029 | 0.60 (0.38–0.96) | 0.035 |
| North Western | 0.41 (0.23–0.75) | 0.004 | 0.41 (0.23–0.73) | 0.003 |
| Southern | 0.74 (0.45–1.24) | 0.254 | 0.76 (0.46–1.25) | 0.273 |
| Western | 0.67 (0.39–1.13) | 0.133 | 0.68 (0.39–1.17) | 0.162 |

*COR: crude odds ratio, AOR: Adjusted odds ratio. We present survey-weighted estimates in this table*

### 3.5 Multivariable logistic regression of adequate ANC on LBW

While adjusting for the number of children ever born, maternal employment status, sex of a child, type of toilet facility, stunting, place of residence and province, attending 4–7 ANC visits as compared to attending 0–3 visits, reduced the odds of low birthweight by 43% (AOR = 0.57; 95% CI: 0.42– 0.78; p < 0.001). Mothers who attended eight or more ANC visits also had significantly reduced odds of having LBW infants (AOR = 0.51; 95% CI: 0.27–0.99; p = 0.047) (Table 2).

An increasing number of children ever born was associated with lower odds of LBW, with each additional child reducing the odds by approximately 10% (AOR = 0.90; 95% CI: 0.84–0.97; p = 0.005). Maternal employment was also protective, as employed mothers had 25% lower odds of delivering a low-birth-weight infant compared with unemployed mothers (AOR = 0.75; 95% CI: 0.57–0.97; p = 0.031). Furthermore, female infants had significantly higher odds of being born with LBW compared with male infants (AOR = 1.32; 95% CI: 1.03–1.69; p = 0.028). Children from stunted mothers had increased odds of LBW relative to non-stunted children (AOR = 1.49; 95% CI: 1.06–2.10; p = 0.021). In addition, rural residence was associated with higher odds of LBW in the crude analysis; however, this association was attenuated and no longer statistically significant after adjustment (AOR = 1.33; 95% CI: 0.98–1.81; p = 0.071). Type of toilet facility was not independently associated with LBW in the adjusted model. In addition, geographic variation in LBW persisted after adjustment. Compared with Central Province, significantly lower odds of LBW were observed in Eastern, Luapula, Northern, and North Western provinces, with the strongest association observed in North Western Province (AOR = 0.41; 95% CI: 0.23–0.73; p = 0.003).

### 3.6 Adjusted predicted probability of LBW by number of ANC visits

The predicted probability of LBW declines steadily as the number of ANC visits increases, indicating a clear inverse gradient (Fig 4). At very low ANC utilization (0–1 visits), the predicted probability of LBW is highest, with wide confidence intervals reflecting greater uncertainty due to fewer observations. As ANC visits increase, the predicted probability decreases sharply through approximately 4–6 visits, after which the decline becomes more gradual. Beyond 8 ANC visits, the predicted probability of LBW continues to decrease but at a slower rate, approaching a low plateau at higher visit numbers. The confidence intervals narrow with increasing ANC visits, suggesting greater precision in the estimated probabilities at higher levels of ANC utilization. Overall, the figure illustrates a dose–response pattern, whereby increasing ANC attendance is associated with progressively lower predicted probabilities of LBW across the observed range of ANC visits.

**Figure 4:**
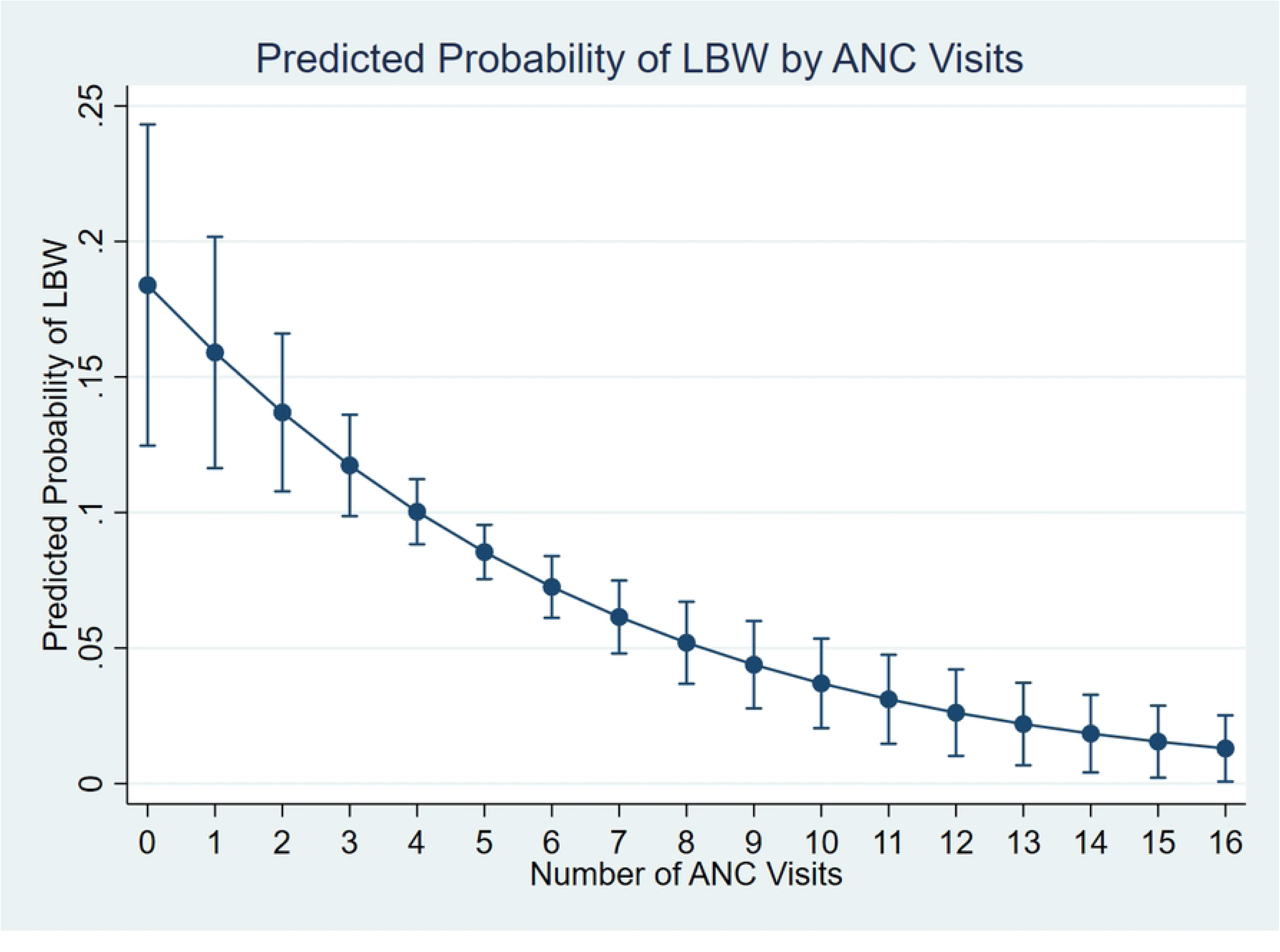
Adjusted probability of LBW by ANC utilization. These probabilities were estimated from survey-weighted multivariable logistic regression whose primary exposure was number of ANC visits which was treated as a count and adjusted for children ever born, maternal employment status, sex of a child, type of toilet facility, stunting, place of residence and province

## 4. Discussion

This study set out to examine the effect of adequate ANC visits on LBW in Zambia. Overall, the findings indicate that the trends of LBW did not vividly decrease with an increase in adequate ANC visits which suggest that there could be other factors which influence LBW besides adequate ANC visits. The Rao-Scott chi-squared test found an association between adequate ANC visits and LBW. In univariable and multivariable logistic regression adequate ANC visits reduced the chances of LBW. These key findings are discussed in detail below in relation to existing literature and their implications for maternal and newborn health policy in Zambia.

### 4.1 Spatial and temporal patterns of adequate ANC visits and LBW

The marked spatial and temporal heterogeneity in LBW prevalence observed over the years and across provinces reflects persistent and evolving inequalities in maternal and newborn health in Zambia. At the national level, LBW prevalence showed relatively modest fluctuations over time, with periods of stabilization coinciding with increases in ANC utilization, particularly following the recovery in ANC utilization after 2013. However, this aggregate national pattern masks substantial subnational variation. Provinces such as Luapula, Western, and Northern, which recorded relatively high LBW prevalence in the early survey years, have historically been characterized by higher levels of poverty, food insecurity, and limited access to quality maternal health services, factors that are well documented contributors to adverse birth outcomes [25,26]. The substantial reductions observed in Luapula and North-Western provinces following the early 2000s may indicate improvements in maternal health service utilization, nutritional interventions, or broader socioeconomic conditions, consistent with regional evidence showing that expanded ANC coverage and maternal nutrition programs can reduce LBW risk over time [27]. However, the resurgence of higher LBW prevalence in recent survey years in Central, Lusaka, Southern, Northern, and Western provinces is concerning and suggests the influence of emerging or unresolved risk factors, including urban–rural disparities, uneven quality of ANC, rising maternal metabolic conditions, and health system disruptions. Similar patterns of stalled or reversed progress in LBW reduction have been reported in other low and middle-income countries undergoing rapid demographic and epidemiological transitions [28]. Collectively, these findings emphasize that progress in reducing LBW has been uneven across space and time, underscoring the need for province-specific, context-responsive strategies that address both longstanding structural determinants and newly emerging risks to maternal and fetal health.

### 4.2 The effect of adequate ANC visits on LBW

This study examined the association between ANC attendance and LBW using recent nationally representative data from the 2024 ZDHS. The findings show that LBW remains a notable public health concern in Zambia, with an overall prevalence of approximately 9%. Both descriptive and regression analyses demonstrated that ANC attendance was strongly associated with LBW, with progressively lower odds and predicted probabilities of LBW observed among women who attended a greater number of ANC visits.

In bivariate analyses, LBW prevalence was highest among mothers who attended 0–3 ANC visits and lowest among those who attended eight or more visits, suggesting a clear gradient across increasing levels of ANC utilization. These patterns persisted in multivariable models, where attendance of 4–7 visits and ≥8 visits was associated with significantly lower odds of LBW compared with inadequate ANC attendance. The adjusted predicted probability plot estimated from an adjusted model which treated ANC as a continuous variable, further illustrated a dose–response pattern, with the probability of LBW declining steadily as the number of ANC visits increased, before plateauing at higher visit numbers. All these results points to the protective effect of ANC on LBW. The observed association between higher ANC attendance and lower odds of LBW is consistent with previous studies conducted in sub-Saharan Africa and other low and middle-income countries. Several studies have found high levels of ANC visits to be protective against LBW [6,10,11,29,30]. The stronger protective association observed among women with 4-7 and eight or more ANC visits aligns with the old and the current WHO recommendation for minimum ANC contacts and provides empirical support for this guideline within the Zambian context.

Adequate ANC has consistently been shown to exert a protective effect against LBW by improving maternal health, nutrition, and early detection of pregnancy-related complications. Through regular ANC contact, pregnant women receive essential interventions such as nutritional counselling and supplementation, screening and management of infections and hypertensive disorders, and timely referral for obstetric care, all of which support optimal fetal growth [9,21]. The protective effect of ANC is particularly evident in malaria-endemic settings, where ANC provides a platform for delivery of intermittent preventive treatment for malaria during pregnancy, a key intervention shown to reduce the risk of LBW [15,16,31]. Moreover, studies incorporating the current WHO recommendation of eight or more ANC contacts suggest a dose–response relationship, with progressively lower odds of LBW observed as the number of ANC visits increases, although the quality and content of care remain critical determinants of benefit [9,32]. Collectively, this evidence supports ANC as an important protective factor against LBW, particularly when visits are timely, frequent, and of adequate

## Strengths and limitations

The strength of this study lies in its use of the latest nationally representative sample allowing for generalizability of the findings to all live births in Zambia. In addition, the study uses both the old and current WHO categorization who allows for comparison of our findings with other studies for credibility. The main limitation of the study is in the way the LBW was collected. Low birthweight was collected from the under-five cards and/or from mother’s recall. Maternal recall of birthweight may suffer from recall bias; however, we mitigated this by including only the most recent birth.

## Conclusion

We determined the effect of Antenatal care visits on Low birthweight using the 2024 Zambian demographic and health survey. We found adequate Antenatal care visits to be protective against low birthweight after controlling for known confounders which included the number of children ever born, maternal employment status, sex of a child, type of toilet facility, stunting, place of residence and province. The findings underscore the importance of ensuring that pregnant women not only access antenatal care but also achieve adequate and sustained attendance throughout pregnancy. Strengthening strategies that promote early initiation and continuity of antenatal care, alongside efforts to improve the quality and content of services delivered, may contribute to reductions in low birth weight. Addressing broader socioeconomic and nutritional determinants of maternal health is also essential to achieving meaningful and equitable improvements in birth outcomes. Overall, reducing the burden of low birth weight in Zambia will require integrated maternal and child health interventions that combine health service delivery with social and nutritional support, particularly for women at higher risk.

## Data Availability

Permission to use the dataset can be acquired from ICF Macro, and the dataset named ZMBR81DTA is available for download at https://www.dhsprogram.com/data

